# Understanding and overcoming innate and acquired MAPK-inhibition resistance in anaplastic thyroid cancer

**DOI:** 10.1101/2024.12.04.24318267

**Authors:** Peter YF Zeng, Jalna Meens, Harrison Pan, Matthew J. Cecchini, Laura Jarycki, Sarah B. Ryan, Alice E. Dawson, Amir Karimi, Mushfiq H. Shaikh, David A. Palma, Eric Winquist, Laksman Gunaratnam, Joe S. Mymryk, John W. Barrett, Paul C. Boutros, Laurie Ailles, Anthony C. Nichols

## Abstract

Anaplastic thyroid cancer (ATC) is one of the most lethal human cancers, with some patients succumbing to the disease within weeks of diagnosis. Although a subset of patients with ATC with BRAF^V600E^ mutation respond to the monomeric type I RAF inhibitor (RAFi) dabrafenib in combination with MEK inhibitor (MEKi) trametinib, most rapidly develop adaptive or acquired resistance. These patients, along with those who do not harbor the BRAF^V600E^ alteration, have limited treatment options. To understand the mechanism of resistance to dabrafenib and trametinib, we utilized multi-region whole genome, high-coverage whole exome and single nuclei RNA-sequencing of ATC patient tumours to unravel genomic, transcriptomic, and microenvironmental evolution during type I RAFi and MEKi therapy. Single-cell nuclei RNA sequencing of matched primary and resistant ATC patient tumours identified reactivation of the MAPK-pathway, along with immunosuppressive macrophage proliferation, underlying the development of acquired resistance. Our translational genomics led us that hypothesize that type II RAFi, which inhibit both RAF monomers and dimers, can be efficacious in overcoming treatment resistance. Screening of a panel of type II RAFi revealed that ATC cell lines are exquisitely sensitive to the type II RAFi, naporafenib, by inhibiting EphA2-mediated MAPK-signaling. We further demonstrated that naporafenib, in combination with the MEKi trametinib, can durably and robustly overcome both innate and acquired treatment resistance to dabrafenib and trametinib using ATC cell lines and patient-derived xenograft models. Finally, we describe a novel mechanism of acquired resistance to type II RAFi and MEKi through compensatory mutations in *MAST1*. Taken together, our work using translational and functional genomics has unraveled the differential mechanisms of treatment resistance to type I and type II RAFi in combination with trametinib and rationalizes the clinical investigation of type II RAFi in the setting of thyroid cancer.

## Introduction

Somatic alterations in the MAPK-RAS pathway are found in over 70% of human cancers^1^. For example, activating mutations in the *BRAF* gene exemplified by the prototypical *BRAF*^V600E^ alteration are observed in more than 70% of melanomas, 30-70% of thyroid cancers, and 10% of colorectal cancers. *BRAF^V^*^600^*^E^* is a clinically actionable alteration as the RAF inhibitor (RAFi) and MEK inhibitor (MEKi) combination dabrafenib and trametinib FDA-approved tissue agnostically^2^. Nevertheless, response rates differ greatly between individual cancer types, suggestive of tissue-specific mechanisms of resistance.

Thyroid cancers serve as an intriguing example of such differential response to MAPK-inhibition. The thyroid is the most common tissue of origin in endocrine malignancy^3^. Differentiated thyroid cancers (DTC) such as papillary thyroid cancer and follicular thyroid cancer (FTC) make up over 90% of all thyroid cancers and are indolent. In contrast, anaplastic thyroid cancer (ATC) accounts for just 1.3% of thyroid cancers but is one of the most lethal human malignancies^4^, with a median survival of 3 – 5 months. Indeed, some patients succumb to the disease within days or weeks of diagnosis. Although TP53 is the most frequently mutated gene in ATC, 40-80% of ATC tumours have alterations in at least one of the members of the RAS-MAPK pathway, such as *BRAF*, *NRAS*, and *PIK3CA*^5–8^. The phase II single-arm, open-label ROAR trial that treated *BRAF*^V600E^ patients with dabrafenib (type I RAFi) and trametinib (MEKi) reported an impressive response rate of 56% and a median overall survival of 86 weeks - a dramatic improvement from historical survival times^9^. These results underscore the importance of inhibition of the MAPK pathway as a therapeutic strategy in ATC. Intriguingly, these response rates are also in stark contrast to the ∼30% response rate seen in the less aggressive DTCs^10^. Unfortunately, almost all ATC patients rapidly developed resistance to type I RAFi/MEKi and have limited further treatment options^4^. Although compensatory alterations in oncogenic RAS^11^, HER3 activation^12^, and MET amplifications^13^ have been linked with resistance to type I RAFi, our understanding of the mechanisms of treatment resistance to RAFi and MEKi in ATC remains incomplete. Furthermore, patients with *BRAF* wildtype tumours have very few treatment options, underscoring the urgent need to mechanistically understand innate and acquired resistance mechanisms to develop more efficacious treatment alternatives. Recently, preclinical and clinical development of type II RAFi have found that they inhibit RAF dimers more effectively and avoid paradoxical activation^2,14^, making them suitable for a broader range of RAF and RAS-driven cancers. Nevertheless, the activity of these compounds in ATC has not been explored.

To address this gap, we sought to understand the mechanisms of innate and acquired resistance to RAFi in ATC. We began with deep molecular phenotyping of genomic, transcriptomic, and cellular evolution of an ATC patient (ATCWGS42) who developed acquired resistance to type I RAFi and MEKi resistance using multi-region whole genome sequencing (WGS), high-coverage whole exome sequencing (WES), and single nucleus RNA-sequencing. Subclonal reconstruction identified MAPK and PI3K-AKT reactivation from an early-branching low-frequency subclone present in the baseline tumour. We further showed that a type II RAFi inhibitor, naporafenib, in combination with trametinib is more efficacious than other RAFi in preclinical models of ATC. In addition, we demonstrate in patient-derived xenograft models of ATC, including one from patient ATCWGS42, that naporafenib and trametinib can overcome innate and acquired resistance to MAPK-inhibition. Mechanistically, naporafenib inhibits EphA2 S897 hyperphosphorylation enriched in thyroid and colorectal cancer and thus co-represses both MAPK and PI3K-AKT signalling. Finally, we demonstrate that compensatory *MAST1* alteration serves as a novel mechanism of acquired resistance to naporafenib. Taken together, our results reveal the mechanisms of innate and acquired resistance to MAPK-inhibition in lethal MAPK-driven cancer and rationalize the clinical investigation of naporafenib in MAPK-cancers that are unresponsive to dabrafenib and trametinib.

## Results

### Genomic, transcriptomic, and microenvironmental evolution during RAFi and MEKi treatment in ATC patient ATCWGS42

We studied an ATC patient with BRAF^V600E^-mutant tumour (ATCWGS42, **Figure 1A**, detailed clinical description can be found in **Methods**) who was treated with the RAFi and MEKi combination, vemurafenib and cobimetinib. The patient demonstrated complete response to vemurafenib and cobimetinib for five months before disease progression, underscoring the rapid occurrence of resistance to type I RAFi and MEKi combination typically seen in patients with ATC. To understand genomic and nongenomic factors that underlie rapid resistance to RAFi and MEKi in ATC, we molecularly dissected the ATCWGS42 tumours pre- and post-RAFi and MEKi treatment (**Figure 1B**).

**Figure 1:**
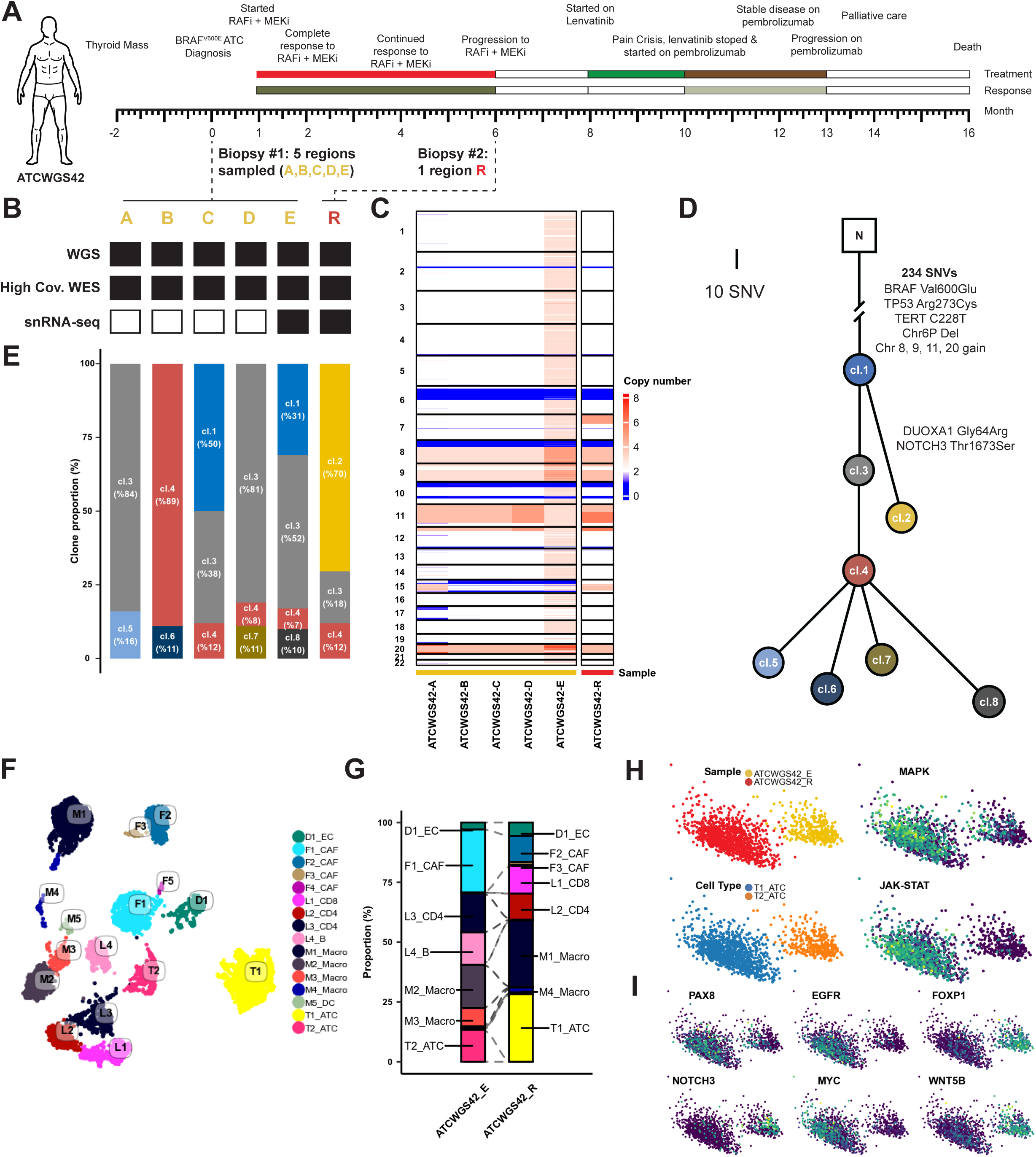
Genomic, transcriptomic, and microenvironmental evolution of ATCWGS42 following RAFi and MEKi treatment. **A)** ATCWGS-42 was a male patient diagnosed with BRAF^V600E^ anaplastic thyroid cancer (ATC). He demonstrated complete response to the RAFi and MEKi combination vemurafenib and cobimetinib. After 6 months, he progressed. Detailed clinical history can be found in **Methods**. Five distinct tumour regions of the treatment-naïve primary tumour and one region of the tumour upon progression were biopsied. **B)** Details of molecular data available from patient. Black means present while white denotes absent. **C)** Copy number alteration landscape across each tumour region. **D)** Subclonal reconstruction of the tumour genomic evolution. The length of the line is proportional to the number of SNV changes, except for between normal and clone 1. Cl.: clone. N: normal **E)** Cancer cell clonal fraction of each subclone, identified in subclonal reconstruction, in each tumour region. **F)** UMAP visualization of cell states identified using single nucleus RNA-sequencing (snRNA-seq) and annotated with finely annotated (level 4) cell type labels. F: fibroblast population. M: myeloid population. L: lymphocyte population. T: tumour. D: endothelial population. **G)** Proportion of cell states within each tumour region. F: fibroblast population. M: myeloid population. L: lymphocyte population. D: endothelial population. T: tumour. EC: endothelial cell. CAF: cancer associated fibroblast. **H)** PCA visualization of ATC tumour cell states, sample, PROGENy inferred MAPK and JAK-STAT pathway output activity scores. **I)** PCA visualization of ATC tumour cell transcript abundance of *PAX8, EGFR, FOXP1, NOTCH3, MYC,* and *WNT5B*.

We performed whole genome sequencing (WGS) and high-coverage whole exome sequencing (WES) of 5 spatially distinct regions of the primary pre-treatment tumour (ATCWGS42-A to ATCWGS42-E) and one post-treatment tumour (ATCWGS42-R). Subclonal reconstruction identified truncal alterations BRAF^V600E^, *TERT* C228T promoter alteration, and multiple chromosomal copy number alterations, including chromosome 6p loss and chromosomes 8, 9, 11, and 20 gain (**Figure 1C**, **D**). We identified a subclone (clone 2) enriched in ATCWGS42-R that makes up more than 70% of the tumour profiled at RAFi/MEKi resistance, suggestive of clonal sweep. (**Figure 1E**). Clone 2 exhibits somatic alterations in *NOTCH3* and *DUOXA1* and was inferred to have branched early from clone 1 (**Figure 1D**). Its early divergence and presence in the primary pre-treatment tumour were further supported by an identical *NOTCH3* alteration in one read within the high-coverage WES of region ATCWGS42-A. To the best of our knowledge, these alterations have not been linked to acquired resistance to dabrafenib and trametinib previously. We did not identify compensatory mutations in other members of the RAS-MAPK pathway, as past studies in melanoma and thyroid cancers have reported^11,14–18^. Nevertheless, the high clonal proportion of clone 2 in the post-treatment ATCWGS42-R tumour suggested that genomic and/or nongenomic features in this clone may offer evolutionary advantages in resistance to MAPK inhibition.

To characterize the transcriptional and tumour microenvironment changes following RAFi and MEKi therapy, we performed snRNA-seq of two tumour components, the pre-treatment (ATCWGS42-E) and the post-treatment tumour (ATCWGS42-R). After stringent quality control to remove low-quality cells and doublets, we clustered the remaining 6367 high-quality cells using the Leiden algorithm^19^ to reveal 16 cell clusters. Manual hierarchical iterative annotation revealed cell types, including two ATC tumour populations, CD4 T-cells, CD8 T-cells, and M2 TAMs (**Figure 1F**). The ATC tumour cell annotation is supported by inferred copy-number states (**Supplementary Figure 1A, B**). In the post-treatment ATCWGS42-R tumour, the vast majority of the macrophage population was annotated as M1_macrophages. The M1_macrophage populations express a high abundance of CD163 and thus are anti-inflammatory CD163 TAMs (**Figure 1G**, **Supplementary Figure 1E**) concordant with previous studies of melanoma patients who develop resistance to dabrafenib^20,21^, suggesting that strategies targeting these populations may relieve the immunosuppressive TME and prolong treatment response.

We next characterized transcriptional changes in the tumour cell populations underlying acquired resistance. ATC cells in the pre-treatment and post-treatment tumours were transcriptionally distinct (**Figure 1H**). Furthermore, the post-treatment resistance tumour showed increased inferred activity of both MAPK and JAK-STAT pathways through PROGENy^22^, suggesting MAPK pathway reactivation. Intriguingly, this MAPK activation is accompanied by loss of expression in NOTCH-pathway genes *NOTCH3*, *FOXP1*, and *WNT5B* and increased *EGFR*, *MYC*, and *PAX8* expression (**Figure 1I**). The decreased abundance of *NOTCH3* in post-treatment tumour cells suggests that the *NOTCH3* alteration in clone 2 identified using WGS and WES may be a loss of function alteration. Furthermore, feedback activation of EGFR is known to cause acquired resistance to RAFi^23^. An increase in the MAPK output score in the post-treatment sample further supported this notion. High expression of the thyroid differentiation marker and transcription factor *PAX8* (**Figure 1I**) served as evidence of RAFi/MEKi induced redifferentiation in ATC^24,25^. Taken together, our detailed genomic and single-cell transcriptomic analysis highlighted the genomic and tumour microenvironmental changes underlying acquired resistance to RAFi/MEKi and strongly underscored reactivation of the MAPK-pathway in the development of resistance to RAFi and MEKi.

### Naporafenib can overcome intrinsic treatment resistance in thyroid cancer

Reactivation of the MAPK pathway following acquired resistance to RAFi and MEKi suggests that vertical inhibition of the pathway may both lead to a more durable response in treatment-naïve patients and be efficacious in combatting acquired resistance. Type II RAFi inhibit ARAF, BRAF and/or CRAF, making them more effective against RAF dimers and broadly applicable in tumours driven by RAS mutations or RAF dimerization led MAPK-activated cancers. They are efficacious in preclinical models of RAF- and RAS-driven melanoma and a subset of patients in other cancer types^26–35^. However, their efficacy in thyroid cancers remains largely unexplored. We screened a panel of 9 thyroid cancer cell lines (8 ATC and 1 PTC cell line K1) with dabrafenib or one of four type II RAFi (belvarafenib, lifirafenib, naporafenib, and MLN2480). No thyroid cell lines were sensitive (defined as IC50 < 10 µM) to dabrafenib (**Figure 2A**), data supported by previous work. The type II RAFi belvarafenib, MLN2480, and lifirafenib inhibited growth in a minority of the screened thyroid cell lines. As a positive control, the BRAF^V600E^ mutant melanoma cell line A375 was sensitive to all tested RAFi. Strikingly, single agent naporafenib genotype-agnostically inhibited the growth of 8/9 thyroid cancer cell lines, including BRAF^V600E^–mutant (8505C, BHT101, K1, SW1736), *NRAS*-mutant (ASH3, KMH2), and *BRAF NRAS*-wildtype cell lines (THJ29T, UHTh7).

**Figure 2:**
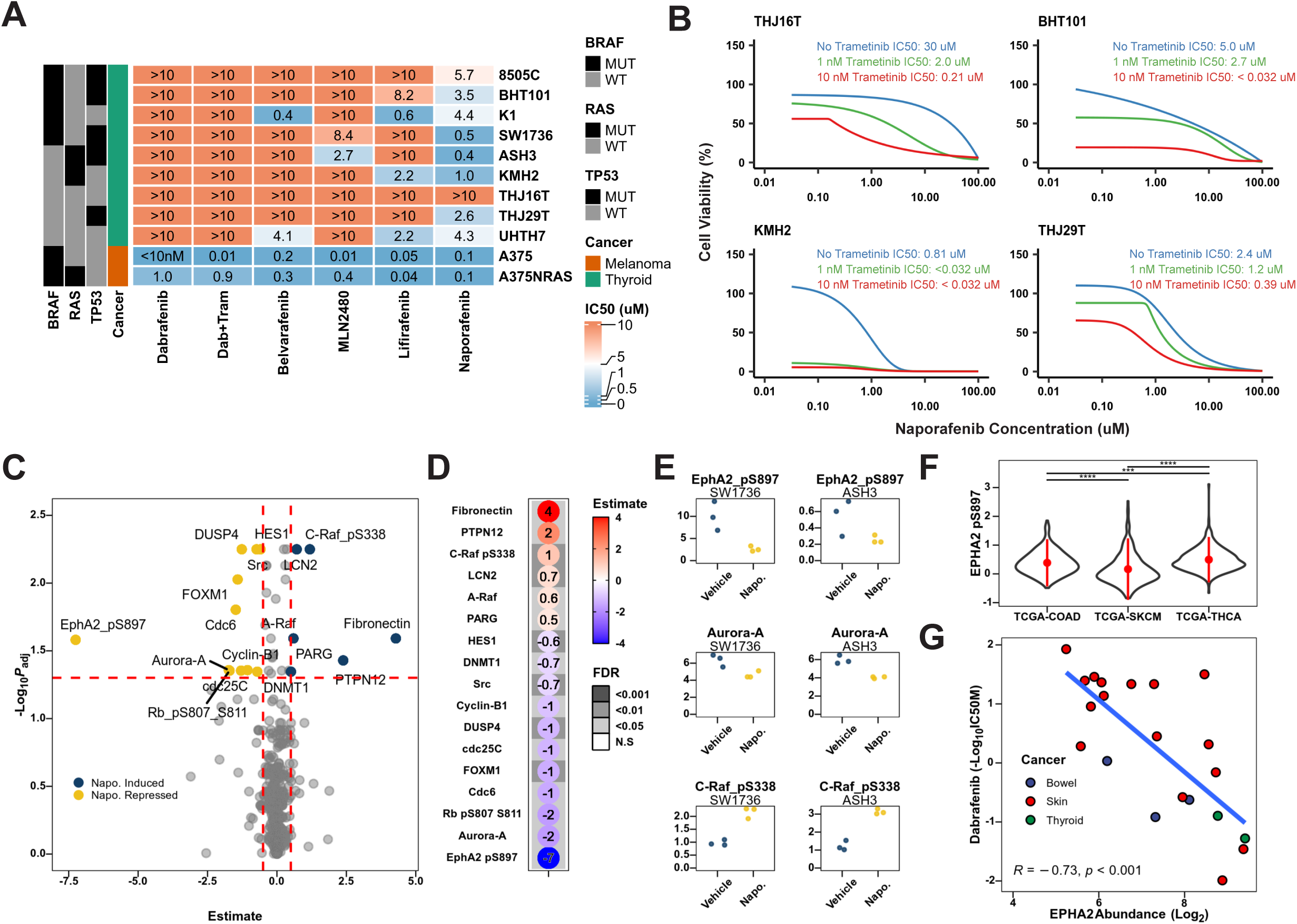
Naporafenib overcomes thyroid cancer resistance to RAFi by repressing EphA2 phosphorylation. **A)** In a panel of 9 thyroid cancer cell lines, the melanoma cell line A375, and its isogeneic A375-NRAS Q61K cell line, naporafenib was found to be efficacious. IC50: half maximal inhibitory concentration. MUT: mutant. WT: wildtype. **B)** The addition of the MEK inhibitor trametinib at nanomolar concentration further sensitizes thyroid cancer cell lines to naporafenib, including the previously resistant cell line THJ16T. **C)** Volcano plot of reverse-phase protein array of cell lines SW1736 and ASH3 treated with either vehicle or naporafenib for 24 hours. linear regression model. Estimate and Benjamini-Hochberg adjusted *P* value from linear regression model, with the following terms: treatment (vehicle or naporafenib), cell line (SW1736 or ASH3), and their interaction term. Proteins/phosphoproteins in yellow are repressed by naporafenib while genes in dark blue are induced by naporafenib, respectively. Horizontal red lines indicate adjusted *P*-value cutoff of 0.05, while vertical red lines indicate effect size cut off of +0.5 or -0.5. **D)** Dotplot of protein/phosphoproteins found to pass both the adjusted-*P* value and effect size cutoff. Shade indicates the adjusted *P*-values while the color of the dot and text inside the circle indicates the estimate. **E)** Abundance of EphA2 pS897, Aurora-A, and CRAF pS338 in SW1736 and ASH3. **F)** EphA pS897 levels in the Cancer Genome Atlas (TCGA) reverse phase protein array of colorectal cancer (TCGA-COAD), skin cutaneous melanoma (TCGA-SKCM), and thyroid cancer (TCGA-THCA) cohorts. *P*-values from Benjamini-Hochberg adjusted Wilcoxon’s rank sum test. ***: FDR < 0.001, ****: FDR < 0.0001. **G)** Scatter plot of EphA2 mRNA abundance and dabrafenib - Log_10_ IC50 in bowel, skin, and thyroid cancer cell lines in the Cancer Cell Line Encyclopedia with BRAF^V600E^. *P*-value and R coefficient from Pearson’s correlation.

Naporafenib is an ARAF-sparing type II RAFi that potently inhibits monomeric or dimeric BRAF and CRAF^31,32^. Naporafenib, in combination with MEKi trametinib, demonstrated a disease control rate of 73.3% in heavily pretreated *BRAF* or *NRAS* mutant non-small cell lung cancer and melanoma patients^36^. We next tested whether the addition of trametinib further sensitized thyroid cell lines to naporafenib. Three different doses of trametinib (0, 1, or 10 nM) were tested in combination with a dose range of 32 nM to 100µM of naporafenib. Importantly, the addition of trametinib at nanomolar concentrations further sensitized thyroid cell lines to naporafenib (**Figure 2B**), including the previously naporafenib-resistant cell line THJ16T, a BRAF-fusion ATC cell line known for its drug resistance. As no BRAF^V600E^ ATC cell line is exquisitely sensitive to dabrafenib/trametinib (**Figure 2B**), we utilized the dabrafenib/trametinib-sensitive A375 melanoma cell line to test the ability of naporafenib in combating dabrafenib/trametinib resistance *in vitro*. We used both the parental A375 cell line and an isogenic CRISPR-Cas9 edited line possessing clinically relevant NRAS Q61K point mutation identified in melanoma patients with acquired resistance to dabrafenib/trametinib, which are known to be more resistant to dabrafenib/trametinib. Naporafenib was equally effective in inhibiting the growth of both the parental and NRAS mutant A375 lines (**Figure 2A**). These data established that combined naporafenib and trametinib treatment could inhibit the growth of cancer cell lines with acquired dabrafenib/trametinib resistance *in vitro*.

### Naporafenib blunts EphA2-mediated intrinsic RAFi resistance

To mechanistically understand the sensitivity of thyroid cancer cell lines to naporafenib, we performed targeted phosphoproteomics using reverse phase protein arrays (RPPA) of BRAF^V600E^-mutant SW1736 and NRAS-mutant ASH3 cell lines treated with either vehicle or naporafenib for 24 hours. Naporafenib induced increased phosphorylation of CRAF and decreased abundance of Aurora A. Strikingly, naporafenib treatment led to a 7-fold decrease in phosphorylation levels of EphA2 S897 (linear regression, **Figure 2C**-**E**, see **Methods**). EphA2 is a member of the receptor tyrosine kinase (RTK) family erythropoietin-producing hepatocellular (EPH) receptors and impinges on the RAS-MAPK, JAK-STAT, and RAS-PI3K-AKT pathways^37–40^. Importantly, EphA2 is well-established to be a critical mediator of resistance to type I RAFi^41^. In The Cancer Genome Atlas (TCGA), thyroid cancers exhibit higher phosphorylated EphA2 S897 compared to both colorectal cancer and melanoma (FDR < 0.001, Wilcoxon’s test, **Figure 2F**), an effect that is consistent even when restricted just to BRAF^V600E^ mutant tumours (**Supplementary Figure 2A**). In the cancer cell line encyclopedia (CCLE), EphA2 mRNA abundance is also higher in cell lines from bowel or thyroid, compared to those from the skin (**Supplementary Figure 2B**). Further, EphA2 mRNA abundance is strongly correlated with dabrafenib IC50 values from cell lines in the CCLE (*P* < 0.001, Pearson’s Correlation, **Figure 2G**). Collectively, these data established EphA2 as an organ-site-specific mediator of RAFi-resistance, with the highest abundance in thyroid cancers.

### Naporafenib and trametinib overcomes resistance to RAFi and MEKi in ATC PDX model of ATCWGS42

Next, we assessed whether naporafenib is efficacious in patient-derived xenograft (PDX) models of ATC. We generated a treatment naïve PDX model (**Figure 3A**) from the pretreatment tumour from the same patient with the BRAF^V600E^ alteration that we studied in detail (ATCWGS42, **Figure 1**). We treated the PDX model with one of DMSO, dabrafenib, trametinib, naporafenib, dabrafenib + trametinib, or naporafenib + trametinib. Whereas dabrafenib alone failed to control tumour growth, naporafenib monotherapy successfully restrained tumour growth until the endpoint. Furthermore, treatment with the naporafenib and trametinib combination led to a complete response (100% maximal tumour control) in 3/5 mice (**Figure 3B**) and was superior at tumour control compared to the other conditions (*P <* 0.001 vs all other groups, linear mixed-effects model, **Figure 3C**, **D**).

**Figure 3:**
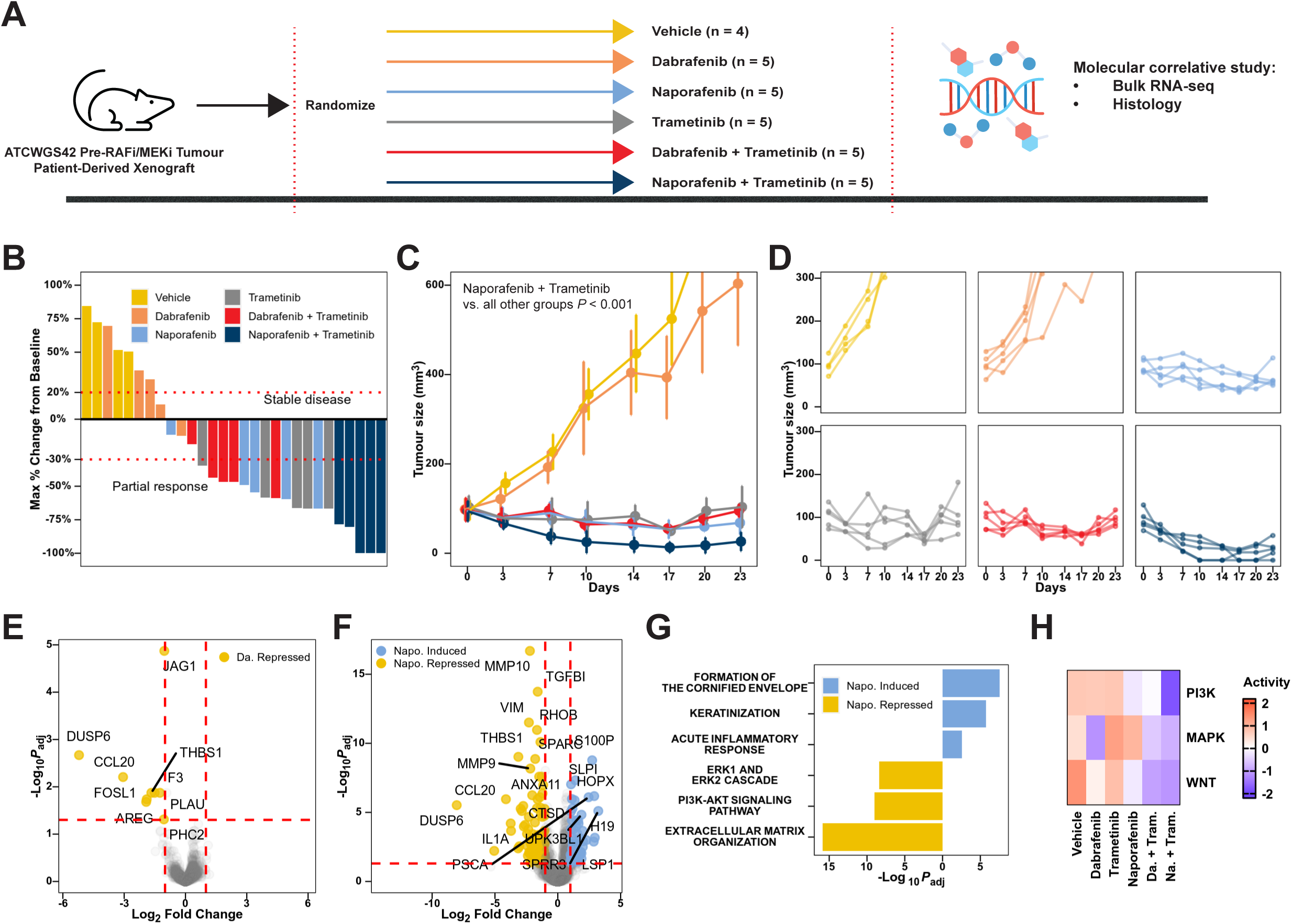
Naporafenib and trametinib induces tumour regression in ATCWGS42-derived xenograft model by co-inhibiting MAPK and PI3K-AKT signalling. **A)** Schematic of experiment. One treatment-naïve region of tumour from ATCWGS42 was used to generate a patient derived xenograft (PDX) model, which was passaged to generate the indicated treatment cohorts. When tumours reached 100mm^3^, mice were randomized into one of six groups: vehicle, dabrafenib, naporafenib, trametinib, dabrafenib and trametinib, and naporafenib and trametinib and treated daily for five days per week. After day 23, all mice were sacrificed. Tumours were collected for molecular correlative analysis using bulk RNA-seq and histology. **B)** Maximal percentage tumour size shrinkage, as defined using RECIST version 1.1. **C,D)** Tumour size plot of each arm grouped (**C**) and spaghetti plot (**D**) showing individual mice. *P*-value from linear mixed-effect model. **E,F)** Volcano plot of transcripts induced and repressed by dabrafenib (**E**) or naporafenib (**F**) treatment. Horizontal red lines indicate adjusted *P*-value cutoff of 0.05, while vertical red lines indicate log_2_ fold change cut off of +1 or -1. *P*-values from DESeq2. **G**) Pathway analysis of transcripts induced and repressed by naporafenib, identified using GProfiler. **H)** Scaled mean pathway output activity of MAPK, PI3K-AKT, and WNT inferred using PROGENy, normalized between groups.

To untangle *in vivo* effects of naporafenib compared to dabrafenib and trametinib, we performed bulk RNA-seq of three tumours from each group at the endpoint. After the removal of mouse reads (see **Methods**), we utilized a multi-factor design to account for the perturbational effects of RAFi (vehicle, dabrafenib, or naporafenib) and MEKi (vehicle or trametinib) and their interactions. Both dabrafenib (**Figure 3E**) and naporafenib (**Figure 3F**) repressed the abundance of a similar subset of transcripts such as *DUSP6*, a negative regulator of MAPK-signalling, suggesting that both dabrafenib and naporafenib repressed MAPK-signalling^42,43^. Furthermore, naporafenib repressed the expression of transcripts critical to epithelial-mesenchymal transition, such as *VIM*^44,45^ (**Figure 3F**). Pairwise comparison between mice treated with naporafenib and trametinib versus dabrafenib and trametinib also identified *VIM* to have higher abundance in the dabrafenib and trametinib group (**Supplementary Figure 2C**). Pathway analysis using GProfiler2^46,47^ (**Figure 3G**) suggested that naporafenib induced the expression of genes related to epithelial function while downregulating ERK and PI3K-AKT signalling. Finally, pathway activity inference (**Figure 3H**) using PROGENy^22^ demonstrated that combined treatment of naporafenib and trametinib uniquely repressed pathway activity of the MAPK, PI3K-AKT, and WNT pathways. These experiments, in sum suggest that naporafenib can overcome innate resistance to MAPK by inhibiting both MAPK and PI3K-AKT signalling, leading to more efficacious tumour control.

### Naporafenib and trametinib overcome acquired resistance to dabrafenib and trametinib

We then assessed the efficacy of naporafenib and trametinib combination therapy in the setting of acquired resistance to dabrafenib and trametinib. We expanded the PDX tumours that were treated with dabrafenib and trametinib (as in **Figure 3** dabrafenib and trametinib group). We implanted ten mice with these tumours and, upon reaching 100mm^3^, began treatment with dabrafenib and trametinib until resistance (**Figure 4A**). Tumour sizes did not shrink but remained steady until day 21 **(Figure 4B**, **C**). On day 33, when the tumours had acquired resistance to dabrafenib and trametinib (mean tumour size reached 250 mm^3^), we randomized the mice to either continue treatment with dabrafenib and trametinib or switch to naporafenib and trametinib. Strikingly, the naporafenib and trametinib combination treatment induced significant tumour regression in dabrafenib and trametinib-resistant tumours (**Figure 4B**, **C**). When all dabrafenib and trametinib-treated mice reached the endpoint by day 64, three mice treated with naporafenib and trametinib were also collected for correlative biomarker analysis. The two mice that remained on naporafenib and trametinib maintained tumour control. From days 95 to 106, the mice went on a forced drug holiday for 11 days (due to institutional policy; see **Figure 4A figure legends**). Post-drug holiday and upon resumption of naporafenib and trametinib treatment, tumour sizes shrunk again before finally reaching the humane endpoint.

**Figure 4:**
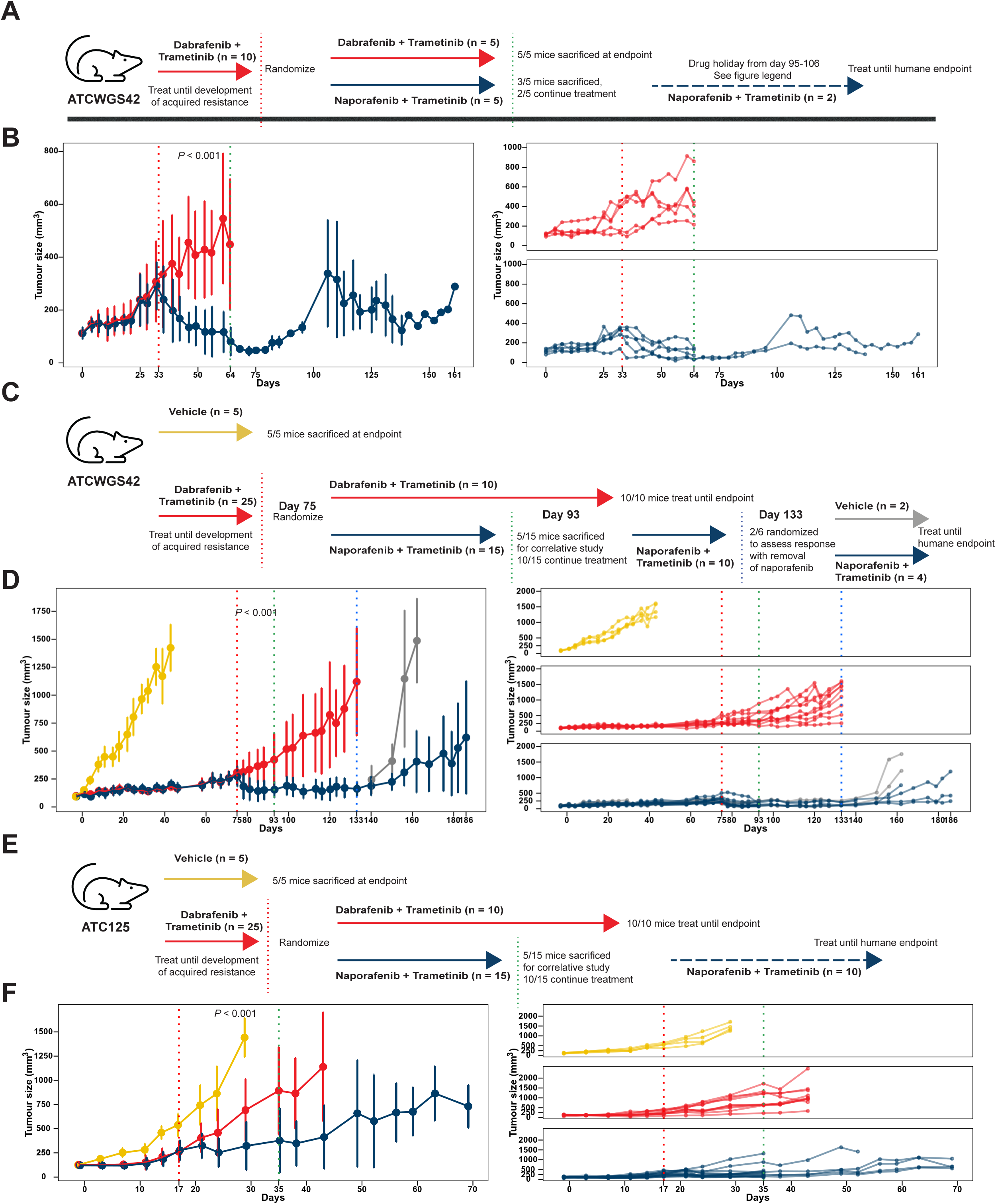
Naporafenib and trametinib overcomes acquired resistance to dabrafenib and trametinib. **A)** Schematic of experiment. The ATCWGS42 PDX model was used to generate 10 mouse tumours. When tumours reached 100mm^3^, mice were treated with dabrafenib and trametinib until tumour size reached 250mm^3^. Mice were then randomized to either continue dabrafenib and trametinib, or switch to naporafenib and trametinib treatment. On day 64 when most mice in the dabrafenib and trametinib group were close to endpoint, all five mice in the dabrafenib and trametinib group and three of five from the naporafenib and trametinib group were sacrificed for correlative studies. Two out of five in the naporafenib continued treatment until they development of resistance or humane endpoint. There was a drug holiday from day 95-106 due to lack of access to the animal facility given institutional mandate. **B)** Tumour size plot of each arm grouped and spaghetti plot showing each individual mouse. *P*-value from linear mixed-effect model comparing naporafenib and trametinib to dabrafenib and trametinib. **C)** Schematic of experiment. The ATCWGS42 PDX model was used to generate 30 mouse tumours. When tumours reached 100mm^3^, mice were treated with vehicle (n=5) or dabrafenib and trametinib (n=25). When the mean tumour size of the group treated with dabrafenib and trametinib reached 250mm^3^, mice were randomized to either continue dabrafenib and trametinib (10 out of 25), or switched to naporafenib and trametinib treatment (15 out of 25). All mice in the dabrafenib and trametinib group were treated until endpoint, and 5 out of 15 mice treated with naporafenib and trametinib were sacrificed at that time. The remaining 10 out of 15 mice naporafenib and trametinib mice continue treatment. On day 133, all the dabrafenib and trametinib group were sacrificed, and 2 of the remaining 6 mice treated with naporafenib and trametinib were randomly selected to vehicle treatment to assess whether removal of treatment results in tumour growth rebound. **D)** Tumour size plot of each arm grouped and spaghetti plot showing each individual mouse for ATCWGS42. *P*-value from linear mixed-effect model comparing naporafenib and trametinib to dabrafenib and trametinib. **E)** Schematic of experiment. A treatment-naïve region of tumour from patient ATC125 was used to establish a PDX model, which was passaged to generate 30 tumours. These were treated in the same way as the ATCWGS42 PDX model above. **F)** Tumour size plot of each arm grouped and spaghetti plot showing each individual mouse for ATC125. *P*-value from linear mixed-effect model comparing naporafenib and trametinib to dabrafenib and trametinib.

We further confirmed these findings by repeating the acquired resistance experiment starting from the treatment-naïve tumour (**Figure 4D**). Mice were treated with either vehicle or dabrafenib and trametinib. On day 75, when the mean tumour size reached 250 mm^3^, mice treated with dabrafenib and trametinib were randomized to either continue dabrafenib and trametinib or switched to naporafenib and trametinib. Strikingly, combined treatment using naporafenib and trametinib led to tumour size regression (*P <* 0.001, linear mixed-effects model, **Figure 4D**). By day 133, average tumour size of mice in the dabrafenib and trametinib group was 1120 mm^3^ and were all sacrificed, while the average tumour size in the naporafenib and trametinib group was 162 mm^3^ (**Figure 4C**, **D**). We tested whether tumour growth relied on continuous naporafenib and trametinib administration by switching two of the remaining six mice treated with naporafenib and trametinib to vehicle treatment (grey group, **Figure 4C**, **D**). Indeed, withdrawal of naporafenib and trametinib led to rapid tumour growth in two mice that both reached endpoint by day 162. Meanwhile, tumour inhibition for three mice out of four mice continuously treated with naporafenib and trametinib continued until day 186, demonstrating the durable and robust response of ATC to naporafenib and trametinib treatment.

Finally, using a PDX-model from the BRAF^V600E^ ATC tumour from a separate ATC patient (ATC125, **Figure 4E**), we found that naporafenib and trametinib combination was also able to control tumour growth more effectively than dabrafenib and trametinib effectively (ATC125, *P <* 0.001, linear mixed-effects model, **Figure 4F**). Taken together, the naporafenib and trametinib combination is efficacious in PDX models of acquired resistance to dabrafenib and trametinib in ATC.

### Acquired resistance to naporafenib and trametinib through compensatory alterations in *MAST1*

Whereas the genomic mechanism of resistance to type I RAFi of *BRAF*^V600E^ is more well-characterized, the mechanisms of resistance towards type II RAFi remain largely unexplored. Compensatory alteration along MAPK pathway members, for example, in *ARAF*, confer resistance to the type II RAFi such as belvarafenib and naporafenib^48^. Nevertheless, additional alternative mechanisms of resistance may exist for naporafenib. To understand the differential mechanism of resistance, we performed high-coverage WES in ATCWGS42 PDX tumours (**Figure 4A**, **B**) at baseline untreated, time of resistance to dabrafenib and trametinib, while responding to naporafenib and trametinib, and finally after acquired resistance to naporafenib after 128 days of naporafenib treatment. The only protein-coding alteration that was significantly enriched in the naporafenib and trametinib resistant PDX tumour was a *MAST1* Ala733Val substitution. This alteration was found in only region E of the original ATCWGS42 tumour at very low frequency (1/493 reads) and slightly enriched in PDX tumours resistant to dabrafenib and trametinib or the tumour responding to naporafenib and trametinib. Strikingly, the VAF for this alteration expanded to 47% of all reads in the tumour that is resistant to naporafenib and trametinib (**Figure 5A**). MAST1 is a member of the CRAF-MEK1 complex and can phosphorylate MEK1 independent of CRAF, leading to CRAF-independent activation of the MAPK-pathway^49^. As naporafenib is hypothesized to bind BRAF and CRAF with similar affinity, compensatory alterations in *MAST1* may thus drive reactivation of MAPK-signalling and tumour proliferation. Indeed, expression of *MAST1* Ala733Val in the ATC cell line BHT101 induced resistance to naporafenib (**Figure 5B**) compared to wildtype *MAST1*, regardless of the presence or absence of trametinib. To the best of our knowledge, this is a novel mechanism of resistance to type II RAFi to reactivate MAPK-signalling (**Figure 5C**).

**Figure 5:**
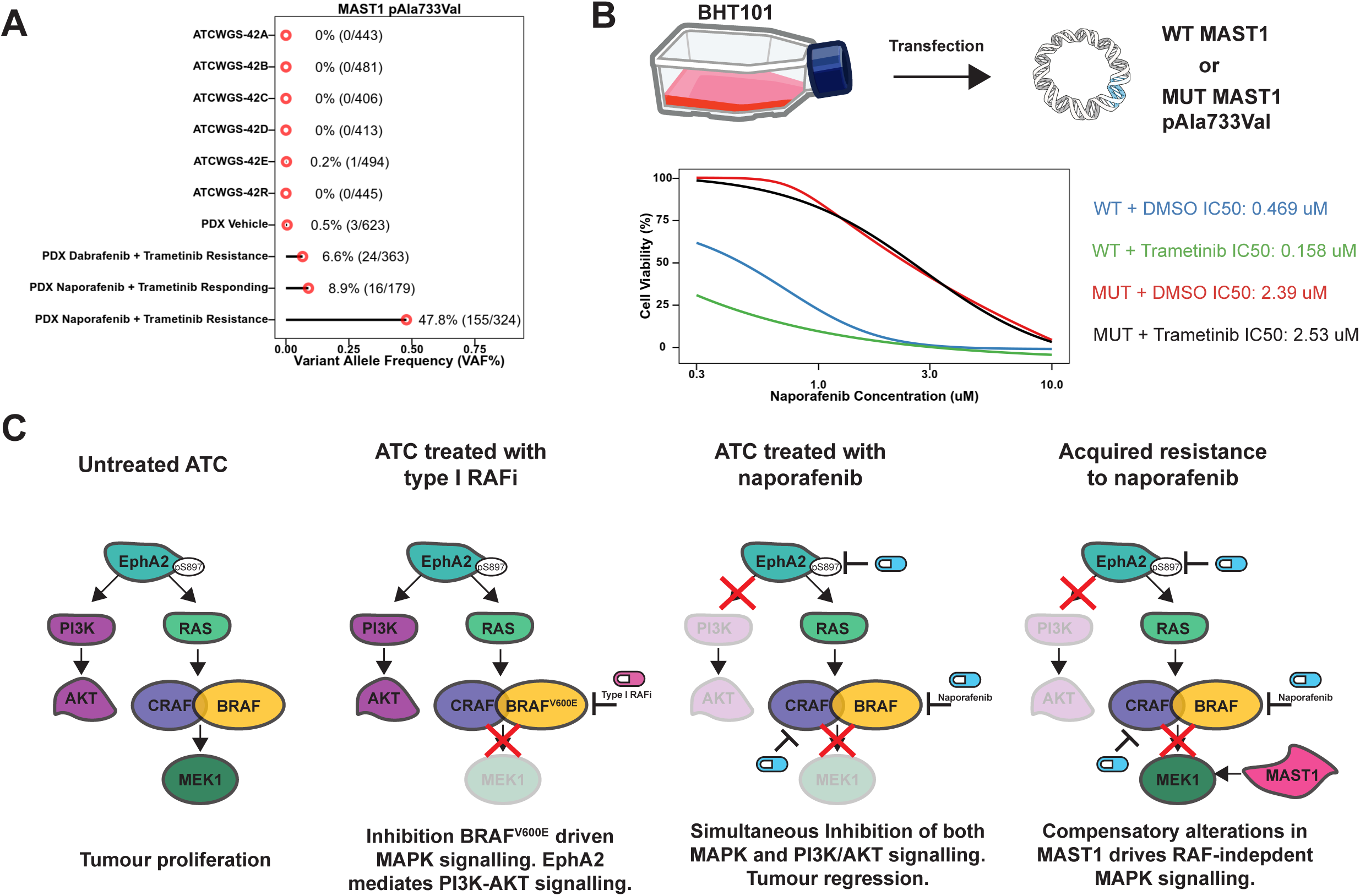
Compensatory alterations in *MAST1* may lead to re-activated MAPK-signalling. **A)** Variant allele frequency of MAST1 Ala733Val alteration in primary tumour (ATCWGS42-A to ATCWGS42-E), post vemurafenib and cobimetinib resistant-tumour (ATCWGS42-R), patient-derived xenograft (PDX) treated with vehicle, PDX model that developed acquired resistance to dabrafenib and trametinib, PDX model that still respond to naporafenib and trametinib, and PDX model that demonstrate acquired resistance to naporafenib and trametinib. Percentage is the total number of reads, while the number in the bracket indicates the number of reference and alternative reads. **B)** IC50 of ATC cell line BHT101 transfected with either wildtype or mutant MAST1 Ala733Val to naporafenib. Each curve represents three biological replicates. **C)** Proposed model of innate and acquired resistance to type I and type II RAF-inhibition in thyroid cancer.

## Discussion

In the current study, we utilize translational and functional multi-omics, paired with *in vitro* and *in vivo* functional studies, to understand and resolve the mechanism of innate and acquired resistance to MAPK-inhibition in lethal thyroid cancer. Further, we demonstrate that the type II RAFi naporafenib can lead to repression of both MAPK and PI3K-AKT signalling models of ATC. In addition, we identify compensatory alterations in MAST1 as a novel mechanism of acquired resistance to type II RAFi inhibition.

Biologically, we identified EphA2 as an important mediator of RAFi resistance that is enriched in thyroid carcinomas and may explain the lower response rate of thyroid cancers towards type I RAFi, compared to melanomas. EphA2 is a receptor tyrosine kinase that activates MAPK signaling by facilitating RAS activation and downstream ERK phosphorylation, as well as PI3K-AKT signaling^51,52^. Thus, our data further underscore the importance of the PI3K-AKT pathway in the aggressivity of ATC. Our recent large-scale genomic and evolutionary analysis of the progression of ATC from the indolent PTC identified an enrichment of *PIK3CA* hotspot alterations^5^, resulting in further dedifferentiation and increased tumour aggressivity^50^. Indeed, inhibition of both MAPK and PI3K-AKT led to prolonged and sustained tumour control in PDX models of ATC^53^. Thus, we add to the mounting evidence that the PI3K-AKT pathway is a critical mediator of response to RAFi.

Methodologically, we demonstrate the advantages of deep molecular phenotyping at multiple time points of a rare cancer type, followed by bedside-to-preclinical modelling to discover and understand mechanisms of treatment resistance in rare cancers. Starting from single patient data, we delineated the genomic, transcriptomic, and microenvironmental evolution during therapy. Insights from led us to identify that MAPK-inhibition using type II RAFi may be efficacious in *in vitro* cell line models of ATC. We further generated PDX models from the same patient and demonstrated that naporafenib and trametinib can more effectively control tumour growth. Finally, we identified additional mechanisms of acquired resistance to naporafenib and trametinib treatment that could lead to additional therapeutic targets. Compared to other MAPK-altered cancer types such as colorectal cancer and melanoma, the tissue-specific mechanism of resistance to RAFi in thyroid cancer remains understudied. The extreme aggressivity of ATC makes it an exceptional case of MAPK-addicted cancer, and as we have demonstrated here, its detailed characterization can illuminate substantial insights to MAPK-directed therapies that may extend to other molecularly similar cancer types. Together, this framework could prove useful for the study of other rare cancers and diseases.

Clinically, our study has important implications for both thyroid cancer and other MAPK-addicted cancers. While ATC is rare, patients suffering from this disease have almost 100% disease-specific mortality, with no standard therapeutic approach^4^. A portion of the 20-50% of patients with tumours harboring BRAF^V600E^ alteration respond to dabrafenib/trametinib^51^, although almost all patients eventually develop resistance^9^. Moreover, no therapy is effective for the ∼50-80% of ATC patients with BRAF-wildtype tumours or those with acquired resistance to RAFi/MEKi. As most patients have rapid disease progression following diagnosis, the wait time for molecular testing (usually 2-4 weeks) precludes some from further treatment. Thus, the development of a genotype-agnostic therapeutic approach allowing for immediate treatment may be especially attractive for ATC. Furthermore, our results may extend to other MAPK-addicted cancer types, such as a subset of colorectal, pancreatic, and lung cancers. Currently, many tissue-agnostic basket trials are enrolling MAPK-altered patients, and our results suggest that it will be important to assess *MAST1* alterations as a putative mechanism of resistance. As clearly demonstrated in both the clinical course and follow-up *in vivo* work of ATCWGS42, treatment with targeted therapies can lead to clonal sweep of subclonal mutations that lead to resistance. Strategies to overcome intratumour heterogeneity are urgently needed to offer more durable responses, for example, through combination treatments with immunotherapy.

In conclusion, our multi-faceted approach, paired with functional studies, has identified and validated novel therapeutic targets to improve patient outcomes for one of the most lethal cancer types. Future work should examine whether our findings apply to BRAF^V600E^ or RAS-mutant tumours from other cancer types such as melanoma and colorectal cancer to evaluate whether the identified effects are thyroid cancer specific or may extend to other MAPK-driven tumour types.

## Experimental Procedures

### Clinical timeline of the patient

This study has been approved by the Western University institutional ethics board (REB#108920) and received written consent from the two ATC patients. ATCWGS42 was a 41-45 year old male who presented with a thyroid mass in December 2018. Ultrasound guided fine needle aspiration (FNA) was highly suspicious of malignancy. In Mar 2019, an attempt at surgical resection was aborted due to extensive laryngotracheal involvement. A total of five spatially distinct tumour sites were biopsied (regions A, B, C, D, E). BRAF^V600E^ mutation status was identified via IonTorrent deep-coverage targeted sequencing, and the patient was started on vemurafenib and cobimetinib in Apr 2019. In May 2019, the patient was confirmed to completely respond to treatment, with complete resolve of neck swelling. Continuous response was seen in Jul 2019. Disease progression was confirmed in Sep 2019. Tracheostomy was performed, and biopsy of the tumour was performed (region R). The patient was started on lenvatinib in Sep 2019, but experienced pain crisis. Lenvatinib was stopped and the patient started pembrolizumab. The patient showed stable disease on Jan 2020. After 8 cycles of pembrolizumab, the patient progressed once again in Mar 2020, and after discussion was transferred to palliative care. The patient unfortunately passed away in June 2020.

### DNA whole genome sequencing (WGS) and whole exome sequencing (WES)

Whole-genome sequencing (WGS) was performed on five spatially distinct regions of the pre-vemurafenib and cobimetinib treatment tumour (regions A, B, C, D, E), one region of the post-treatment tumour (R), and matched normal tissue (blood). Genomic DNA was isolated from snap-frozen tissue and subjected to whole genome sequencing at The Center for Applied Genomics (Hospital for Sick Children, Toronto, ON) or at Beijing Genomic Institute (BGI). High-coverage whole exome-sequencing was performed at GeneSeeq Inc. (Toronto, Ontario, Canada) using IDT xGen exome panel^5,55,56^. Data were aligned to the GRCh38 human reference genome, including available ALT and decoy alleles using BWA-mem (v0.7.17)^57^. Lane-level BAM files were merged using samtools (v1.16.1). GATK4 (v4.3.0) was used to mark duplicate reads and perform base quality recalibration. SNVs were identified using Mutect2 (v4.3.0). Indels and structural variations were called using Manta (v 1.6.0). Allele-specific subclonal copy number alterations were identified using titanCNA (v 1.17.1). Subclonal reconstruction was performed using CONIPHER using copy number calls generated from WGS data and variants generated from high-coverage WES, as previously described^58^. For WES of PDX models, we used a similar approach except that human and mouse reads were first separated using xengsort^59^ (v2.0.1) using default settings before proceeding to alignment of human reads to the human genome and the rest of the workflow.

### Single nucleus-RNA sequencing (snRNA-seq)

We performed single nucleus RNA-sequencing (snRNA-seq) on two regions from ATCWGS42 (ATCWGS42_E and ATCWGS42_R). Fresh tumour tissue was minced into 1-2 mm^3^ pieces on dry ice using a chilled razor blade. The tissue was covered with lysis buffer and transferred to a tube to lyse on ice. The supernatant and pellet were dounced separately and transferred to chilled tubes. Nuclei viability was confirmed by SYBR staining using a Countess Cell Counter (Invitrogen) and collected by centrifugation (800xg for 10 minutes). This process was repeated. Nuclei were then resuspended in 1 ml suspension buffer and passed through a 40 µm Flowmi cell strainer. Nuclei were then loaded onto the Chromium single cell controller (10X Genomics) and used to construct single nucleus 3’ libraries. Sequencing was performed using NovaSeq 6000 sequencer.

Alignment to GRCh38 reference genome, barcode, and unique molecular identifier counting were performed using CellRanger (v6.0.2). Nuclei were quality-controlled and filtered using the following criteria: 1) number of detected genes was above 200) the number of transcript counts detected was above 500. We further removed nuclei whose genes detected, total transcript count, total mitochondrial transcript count, and mitochondrial transcript percentage are 5 median absolute deviation (MAD) above the median. We then logged and normalized transcripts counts per nucleus. We then logged and normalized transcripts counts per nucleus. We integrated the dataset across using harmonypy^54^. In total, we observe a total of 6367 high-quality cells.

To identify cell states, we used the python scanpy (v1.9.3) package to identify highly variable genes (HVG) with a max_mean of 3, a minimum mean of 0.0125, and a minimum dispersion of 0.5 that resulted in 3909 highly-variables genes. Principal component analysis (PCA) was performed on the HVGs. The scanpy neighbors function was used to generate nearest neighborhood graphs. Clustering was performed using the leiden algorithm. UMAP was applied for dimensionality reduction. We performed hierarchical annotations of the identified clusters. In the first level of cell annotations, we used well-characterized cellular markers to annotate cells as epithelial (*FXYD3, EPCAM, ELF3*), immune (*PTPRC, CD53, CORO1A*), stroma (*COL1A3, DCN, MFAP4*), and endothelial (*CLDN5, ECSCR, CLEC14A*), which makes up cell type level 1. We then further annotated cell clusters using marker genes identified using the *scanpy rank_genes_groups* function hierarchically with increasing precision until the finest level (cell type level 4). In total, we identified 1 endothelial population, 2 ATC tumour populations, 4 fibroblast populations, and 9 immune populations (4 lymphoid and 5 myeloid derived).

### In vitro drug screening

All thyroid cancer cell lines were acquired from Japanese Collection of Research Bioresources Cell Bank (JCRB), Deutsche Sammlung von Mikroorganismen und Zellkulturen (DSMZ), University of Uppsala, or the Mayo Clinic and were subjected to regular STR profiling (**Supplementary Table 14B)**. A375 and isogenic CRISPR-Cas9 knock-in *NRAS*-Q61K A375 cell lines were acquired from American Type Culture Collection (ATCC). Cells were plated in 96 well plates. After 24 hours, for screening using only RAFi, cells were treated with the RAFi in 6 replicates to generate 10-point dose response curves ranging from 1.5 nM to 100µM. For experiments assessing the effect of trametinib in addition to naporafenib, three different doses of trametinib (0, 1, or 10 nM) were tested in combination with a dose range of 32 nM to 100µM of naporafenib. After 72 hours, cells were incubated with PrestoBlue, fluorescence readings were taken and mean inhibitory concentrations (IC50) were calculated using the R package nplr (v.0.1-7).

### Functional phosphoproteomics using reverse-phase protein array

Targeted functional phosphoproteomics using reverse-phase protein array (RPPA) was performed at the MD Anderson Cancer Center RPPA core. Briefly, protein extracts were diluted 2-fold in five serial dilutions (undiluted to 1:16) and spotted onto nitrocellulose-coated slides. These extracts are arranged in a 4×12-pin-11×11 subarray that accommodates 1056 serially diluted samples plus replicate vertical controls on each slide. Each protein of interest was detected by probing with a specific antibody, amplifying the signal via a tyramide amplification system, and visualized via DAB colorimetric reaction. Each slide was probed with one antibody. The detection system used was a GenPoint-based staining kit (Agilent). Digital images of slides were obtained by scanning on a Huron TissueScope scanner producing 16-bit TIFF files. Spot intensities from the TIFF files were determined via Array-Pro Analyzer software. Mean net intensities of spots from all 1056 samples were used for curve-fitting for each slide (RPPA Super Position and Concentration Evaluation). Relative protein level for each sample was determined by converging its sample dilution series to one point (EC50) and interpolating in RPPASPACE (MD Anderson).

### Establishment of patient-derived xenograft (PDX) models and treatment

Fresh ATC tumour specimens were received and kept at 4*°* Celsius in PBS until engraftment no later than 24-hours post resection, as we have previously done. Briefly, viable tumour pieces were implanted subcutaneously into the flank regions of *NOD/SCID/IL2Rγ^-/-^ (NSG)* mice. Mice were sacrificed using isoflurane γ anesthesia followed by CO_2_ asphyxiation when tumours reached a diameter of 1–1.5 cm. Tumours were dissociated in culture medium (containing DNASE 1 and collagenase/hyaluronidase) and passaged subcutaneously with five mice per treatment in 1:1 Matrigel/PBS to generate the PDX models. Mice with tumour volumes 80-120 mm^3^ were randomly assigned to groups of 5, and treated daily with vehicle, dabrafenib, trametinib, dabrafenib/trametinib, naporafenib, or naporafenib/trametinib. Dabrafenib (30mg/kg), naporafenib (15mg/kg), and trametinib (0.6mg/kg) were administered via oral gavage 5 days per week for up to 3 weeks. Tumour volume was calculated using the formula (π/6)×length×width×height. Tumour volume comparison was made using linear mixed-effects model, including tumour treatment group and day as fixed effects, and individual mice number as random effects.

### Bulk RNA-sequencing of tumours from PDX models

Tissue was first homogenized with a VWR microhomogenizer, and the homogenate passed through a QiaShredder as previously described^61–63^. Qiagen AllPrep DNA/RNA kits was used to isolate total RNA. RNA was shipped to Geneseeq Technologies (Toronto, ON) for quality control, library preparation, and sequencing^64^. RNA quality was confirmed with Bioanalyzer, and libraries were prepared using a NEB Ultra II Directional mRNA library kit. Samples were then processed using random primers and paired end sequencing using an Illumina HiSeq 2500 yielded 100 million reads/sample. Human and mouse reads were first separated using xengsort^59^ (v2.0.1) using default settings. The human reads were then aligned to the reference genome GRCm38 using STAR (v2.6.1d) two-pass mode, and transcripts abundance were estimated using RSEM (v1.3.1). DESeq2 (v 1.40.2) was used to perform multifactorial-analysis, by including RAFi (vehicle, dabrafenib, or naporafenib), MEKi (vehicle or trametinib), and their interactions terms in the design formula. GProfiler2 (R package gprofiler2 v0.2.2) was used to perform pathways analysis, excluding gene sets with more than 400 or less than 20 genes. PROGENy^22^ as part of the decoupleR (v2.6.0) was used to infer the activity of MAPK, PI3K-AKT, and WNT pathways.

### Transfection of MAST1 expression plasmids

BHT-101 cells were seeded at a density of 2 x 10^6 cells per well in a 6-well dish and were incubated overnight at 37 °C with a supplement of 5% CO_2_. The next day the cells (60-80% confluency) were transfected with the plasmids expressing either WT MAST1 or MAST1 V733A mutation respectively with Lipofectamine^TM^ 3000 Transfection Reagent (Invitrogen) per the manufacturer’s protocol. The cells were then incubated for four hours with the transfection mixture followed by the replacement of the transfection media with complete media. The transfected cells were then placed in the incubator for 72 hours. Blasticidin was added at 5 ug/ml to select successfully transfected cells. Non-transfected parental controls were dead within 48 hours. Drug selection was continued for 1 week to ensure the maximum availability of the transfected cells.

### Drug Treatment and Testing of Resistance

The BHT-101 cell line, transfected with either the plasmid carrying MAST1(wildtype) or MAST1 mutant (A733V) were counted and seeded at 5 x 10^3^ per well in 96 well dishes. The plates were incubated at 37°C for 5 hours. The spent media was removed and fresh media containing either 2 nM trametinib or 0 nM trametinib and various doses of naporafenib (0.15-10uM) was added to the cells. The plates were incubated for up to 72 hours at 37°C. Cell viability was then determined using Presto Blue (Thermo Fisher Scientific, Waltham, MA, USA) viability reagent. Prestoblue was added and the plate was incubated for 60 minutes at 37°C and the fluorescence levels were measured using a Synergy H4 Hybrid Reader (BioTek, Winooski, VT, USA). The cells were excited at 560 nm and the emission measured at 590 nm. Response curves were generated using the R-script nlpr package (v.0.1-7).

### Statistics

All statistical analyses were performed using either R (v4.3.1) or Python (v3.8.17). Wilcoxon test, linear mixed effect models, and Pearson’s correlation were performed using the packages rstatix (v0.7.2), lme4 (v1.1-34), or stats. Unless otherwise indicated in the figure legend or text, correction of multiple comparisons was performed utilizing the Benjamini-Hochberg procedure as previously described. All tests were two-sided.

### Data Availability Statement

All generated bulk RNA-seq, whole exome sequencing, and single nuclei data sets will be made available upon acceptance at EGA. TCGA data was retrieved from the National Cancer Institute Genome Data Commons (https://portal.gdc.cancer.gov/). The cancer cell line encyclopedia was retrieved using CellMinerDB (https://discover.nci.nih.gov/rsconnect/cellminercdb).

## Acknowledgments

We would like to thank all patients and their families who participated in this study.

## Supplementary Figure Legends

**Supplementary Figure 1.**
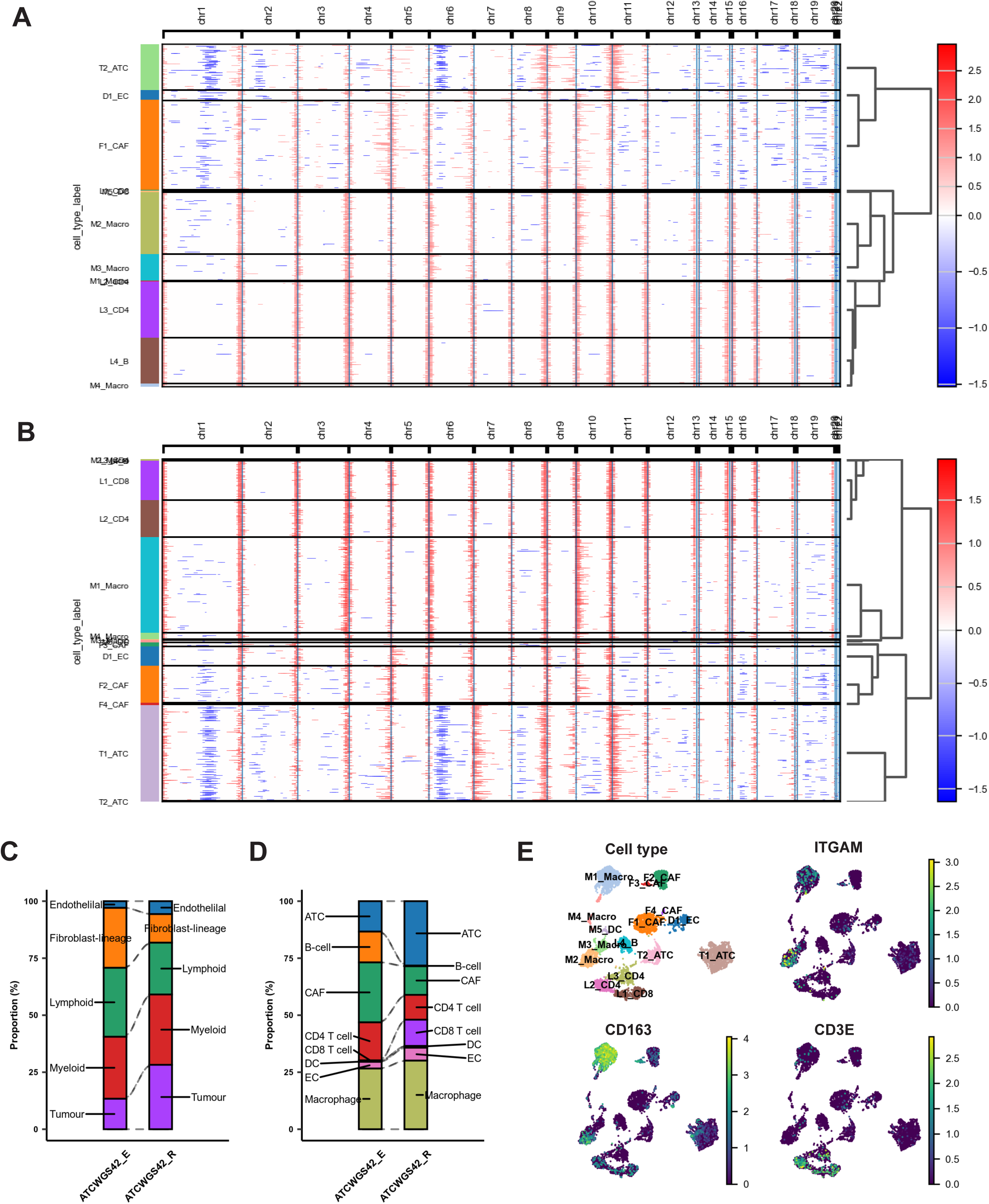
Tumour microenvironment changes in ATCWGS42. **A,B)** Copy number state of cells for ATCWGS42_E (**A**) and ATCWGS42_R (**B**) estimated using inferCNVpy, grouped by their cell type level 4 annotations. **C,D)** Bar plot showing the distribution of cells in each disease and tissue, colored using cell type level 2 (**C**) and level 3 (**D**) annotations. **E)** UMAP of samples colored by cell type, and transcript abundance of *ITGAM*, *CD163*, and *CD3E*.

**Supplementary Figure 2.**
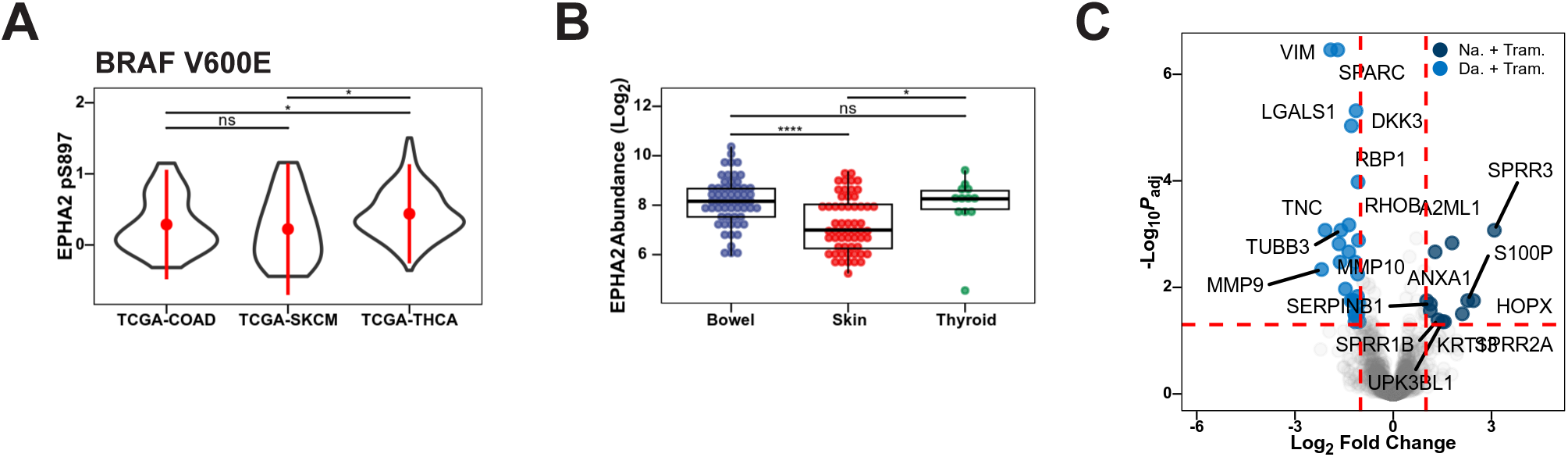
EphA2 may mediate resistance to RAF-inhibition in thyroid cancer. **A)** EphA pS897 levels in the Cancer Genome Atlas (TCGA) reverse phase protein array of colorectal cancer (TCGA-COAD), skin cutaneous melanoma (TCGA-SKCM), and thyroid cancer (TCGA-THCA) cohorts, including only patients with BRAF^V600E^ alteration. *P*-values from Benjamini-Hochberg adjusted Wilcoxon’s rank sum test. *: FDR < 0.05, n.s: FDR > 0.05. **B)** EphA2 mRNA abundance in bowel, skin, and thyroid cancer cell lines in the Cancer Cell Line Encyclopedia. *P*-values from Benjamini-Hochberg adjusted Wilcoxon’s rank sum test. *: FDR < 0.05, ****: < 0.0001, n.s: FDR > 0.05. **C)** Volcano plot of transcript comparison between ATCWGS42 PDX mice treated with dabrafenib and trametinib or naporafenib and trametinib. Horizontal red lines indicate adjusted *P*-value cutoff of 0.05, while vertical red lines indicate log_2_ fold change cutoff of +1 or -1. *P*-values from DESeq2.

